# Treatment of Acute Myocardial Infarction and Cardiogenic Shock: Outcomes of the RECOVER III Post-Approval Study by SCAI Shock Stage

**DOI:** 10.1101/2023.07.25.23293174

**Authors:** Ivan Hanson, Akash Rusia, Andres Palomo, Adam Tawney, Timothy Pow, Simon R. Dixon, Perwaiz Meraj, Eric Sievers, Michael Johnson, David Wohns, Omar Ali, Navin Kapur, Cindy Grines, Daniel Burkhoff, Mark Anderson, Alexandra Lansky, Srihari S. Naidu, Mir B Basir, William O’Neill

## Abstract

**Background:** The Society for Cardiovascular Angiography and Interventions (SCAI) proposed a staging system (A-E) to predict prognosis in cardiogenic shock. Herein, we report clinical outcomes of the RECOVER III study for the first time, according to SCAI shock classification.

**Methods:** RECOVER III is an observational, prospective, multicenter, single-arm, post-approval study of patients with acute myocardial infarction complicated by cardiogenic shock (AMICS) undergoing percutaneous coronary intervention (PCI) with Impella support. Patients enrolled in RECOVER III were assigned a baseline SCAI shock stage. Staging was then repeated within 24 hours after initiation of Impella. Kaplan-Meier survival curve analyses were conducted to assess survival across SCAI shock stages at both timepoints.

**Results:** At baseline assessment, 16.5%, 11.4%, and 72.2% were classified as Stage C, D, and E, respectively. At ≤24 hour assessment, 26.4%, 33.2%, and 40.0% were Stage C, D, and E respectively. Thirty-day survival amongst patients with Stage C, D and E shock at baseline was 59.7%, 56.5% and 42.9%, respectively (p=0.003). Survival amongst patients with Stage C, D and E shock at ≤24 hours was 65.7%, 52.1% and 29.5%, respectively (p<0.001). After multivariate analysis of impact of shock stage classifications at baseline and ≤ 24 hours, only Stage E classification at ≤24 hours was a significant predictor of mortality (OR 4.8, p<0.001).

**Conclusions:** In a real-world cohort of patients with AMICS undergoing PCI with Impella support, only Stage E classification at ≤ 24 hours was significantly predictive of mortality, suggesting that response to therapy may be more important than clinical severity of shock at presentation.

**Clinical Perspective:**

- Many patients with AMICS treated with Impella present in SCAI Stage E shock, but may improve to a more favorable shock stage within 24 hours of their presentation.
- In-hospital and 30-day outcomes correlate with SCAI shock stage at presentation, but the correlation is more robust upon repeat assessment within 24 hours after Impella initiation.
- The SCAI shock staging system may be a useful clinical tool to guide in-hospital management of patients with AMICS, especially within the first 24 hours, and to provide prognostic information to patients and/or their families.

## Introduction

Acute myocardial infarction complicated by cardiogenic shock (AMICS) is a complex syndrome associated with high rates of mortality and morbidity.^1^ While immediate revascularization via percutaneous coronary intervention (PCI) has been the focal point of treatment for the last several decades, ^2, 3^ the optimal strategy for addressing hemodynamic compromise in AMICS has not been definitively established and mortality rates have remained unacceptably high.^4^ Prior studies have shown the intra-aortic balloon pump (IABP) generally provides insufficient support for patients with AMICS and does not reduce mortality.^5^

The Impella 2.5 and CP transcatheter, transvalvular left ventricular pumps (Abiomed, Danvers, MA) are placed in the cardiac catheterization laboratory and can generate up to 4.3 L/min of micro-axial, in-parallel flow, and are indicated in the United States and other countries for the treatment of cardiogenic shock. While the Impella device can have superior hemodynamic effects compared with IABP,^6^ more information is needed about the impact of Impella on survival in different stages of shock severity. The National Cardiogenic Shock Initiative (NCSI) study demonstrated that with protocolized Impella management in the setting of AMICS and a set of “best practices,” a 71% survival to discharge was observed.^7^ Since FDA approval of the Impella CP device in April 2016, an FDA-mandated post-approval study (RECOVER III) has also captured “real-world” outcomes of Impella use in AMICS patients undergoing PCI.

Since 2019, the Society for Cardiovascular Angiography and Interventions (SCAI) shock stages have been essential in not only describing the clinical severity of CS in a given patient at a given timepoint, but also to describe and compare different study populations and their outcomes.^8^ However, it remains unclear at what timepoint SCAI classification has the most prognostic value for early clinical outcome. Herein, we present for the first time the results of the RECOVER III study and further stratify outcomes based on severity of shock using the SCAI shock classification.

## Methods

### Study design and patient population

RECOVER III is an observational, prospective, multicenter, single-arm, FDA-audited post-approval study. Data for RECOVER III were collected within the cVAD registry, a U.S. registry that aims to monitor the post-market performance of Impella heart pumps. The methodology of the cVAD registry (clinicaltrials.gov no. NCT04136392) has been published previously.^9^ This study was conducted in accordance with the Declaration of Helsinki, with IRB/IEC approval at all participating sites. The IRBs waived consent for index admission data. Informed consent was initially required for prospective, post-discharge follow-up data collection, though the study protocol ultimately was revised to grant a waiver of informed consent for retrospective entry of follow-up data for all patients who did not actively decline consent within a 1 year period.

The RECOVER III study assessed outcomes in patients with AMICS who underwent revascularization and received an Impella device before, during, or after revascularization. Patients receiving Impella 2.5, CP, 5.0, or LD heart pumps (providing between 2.5 and 5.3 L/min of flow) were eligible for inclusion. Patients enrolled in the cVAD registry for other indications were excluded. From April 2016 through March 2020, a total of 418 cVAD patients treated at 41 US sites were considered eligible and included in the RECOVER III study. A study flow chart diagram is presented in Supplemental Figure 1.

### Society for Cardiovascular Angiography and Intervention (SCAI) shock stage classification

A chart review was conducted for the purpose of providing supplemental data to perform the SCAI shock stage classification of subjects. As part of the cVAD registry, study records are kept private, and all protected health information (PHI) were de-identified in accordance with US laws. The dataset used to classify shock stage included laboratory values (lactate, pH, creatinine, liver enzymes), clinical exam findings, hemodynamics, vasoactive drug support, mechanical circulatory support, and clinical history (including description of cardiac arrest or unstable arrhythmias) for the separate timepoints.

Patients were assigned a SCAI shock stage by two physicians, each blinded to the classification of the other observer and to the patient outcomes. In the case of discrepant classification, a third blinded physician adjudicated the discrepancy. Staging was performed for two separate timepoints. The first (baseline) SCAI shock stage classification was assigned using the dataset provided from the timepoint nearest, but prior to, Impella placement. The second SCAI shock stage classification was assigned using the dataset from a timepoint within 24 hours after Impella placement. Given that placement of MCS was an inclusion criterion for the RECOVER III study, the minimum SCAI shock stage at baseline and within 24 hours was Stage C. If the appropriate variables were available, the Cardiogenic Shock Working Group-SCAI (CSWG-SCAI) shock staging criteria were used: these have been described in a prior publication.^10^ Shock with cardiac arrest was always classified as Stage E. The CSWG-SCAI shock staging criteria stipulates out of hospital cardiac arrest (OHCA) meets criteria for Stage E based on a sensitivity analysis demonstrating the impact of OHCA on mortality. Given the lack of consistent clinical records describing the duration of the cardiac arrest and post-arrest status, it was decided that in-hospital cardiac arrest also met criteria for Stage E.

### Endpoints and statistical analysis

Clinical characteristics and outcomes were reported for the overall patient population, as well as within SCAI shock stage subgroups (per classification at baseline assessment). The primary endpoint of RECOVER III was survival at 30 days or discharge, whichever was longer. Major adverse cardiac and cerebrovascular events (MACCE), a composite endpoint which comprises death, myocardial infarction, cerebrovascular accident/stroke/transient ischemic attack, and revascularization (including emergent coronary artery bypass grafting), were site reported at time of hospital discharge, 30 days, 90 days, and 1 year.

Kaplan Meier survival curve analyses were conducted to assess survival across shock stage subgroups through 30 days. Regression analyses to identify predictors of mortality (at 30 days or discharge, whichever is longer) were performed using a logistic model. The multivariate analysis of clinical variables was performed using a forward fit stepwise approach. All variables in the univariate analysis were considered, with exception of the variables of the pH and lactate, which were not eligible for the model due to a large number of missing values. Multivariate analysis of the SCAI shock stage classifications at baseline and ≤ 24 hours and their respective associations with early mortality was conducted with each staged treated as an ordinal staged category, with Pre SCAI Stage C and Post SCAI Stage C the reference variables. Statistical analyses were performed using SAS version 9.4 (SAS Institute, Cary, NC). All p values are two-tailed, with p < 0.05 considered statistically significant.

## Results

### SCAI shock stage classification and baseline characteristics

Of 418 patients enrolled in RECOVER III, 413 patients had data available to classify baseline SCAI shock stage prior to Impella placement, and 415 within 24 hours of Impella placement. Of these 413 patients, 68 patients (16.5%) were classified as Stage C, 47 (11.4%) Stage D, and 298 (72.2%) Stage E at baseline assessment. Baseline characteristics of all RECOVER III subjects and within the Stage C, D, and E subgroups (per baseline assessment) are presented in Table 1. Stage C patients were more likely to be white (80.9%) compared to Stage D/E patients (68.1% and 70.1%, respectively). Only 5.9% of Stage C patients were black, compared to Stage D/E patients (12.8% and 13.4%, respectively). Interestingly, LVEF was lower in Stage C patients (22.9%) compared with Stage D/E patients (27.8% and 26.4%, respectively). Baseline lactate in Stage C, D, and E patients was 2.9 ± 1.5, 3.8 ± 2.4, and 9.2 ± 7.6 mmol/L respectively (Table 2). Baseline pH was 7.32 ± 0.06, 7.32 ± 0.07, and 7.12 ± 0.56 in Stage C, D, and E patients, respectively.

**Table 1.**
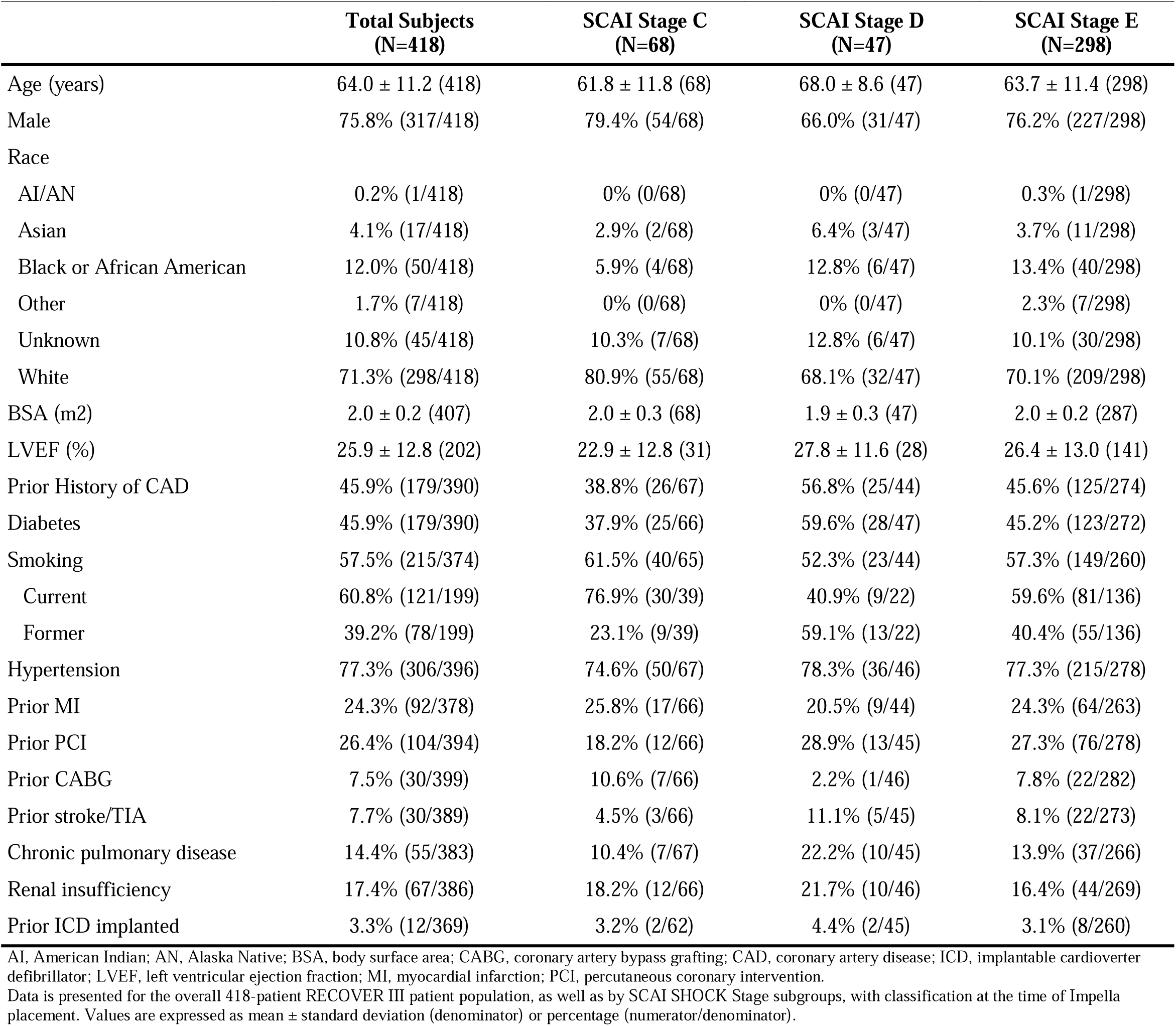
Baseline characteristics, all RECOVER III patients and stratified by baseline SCAI SHOCK Stage.

**Table 2.**
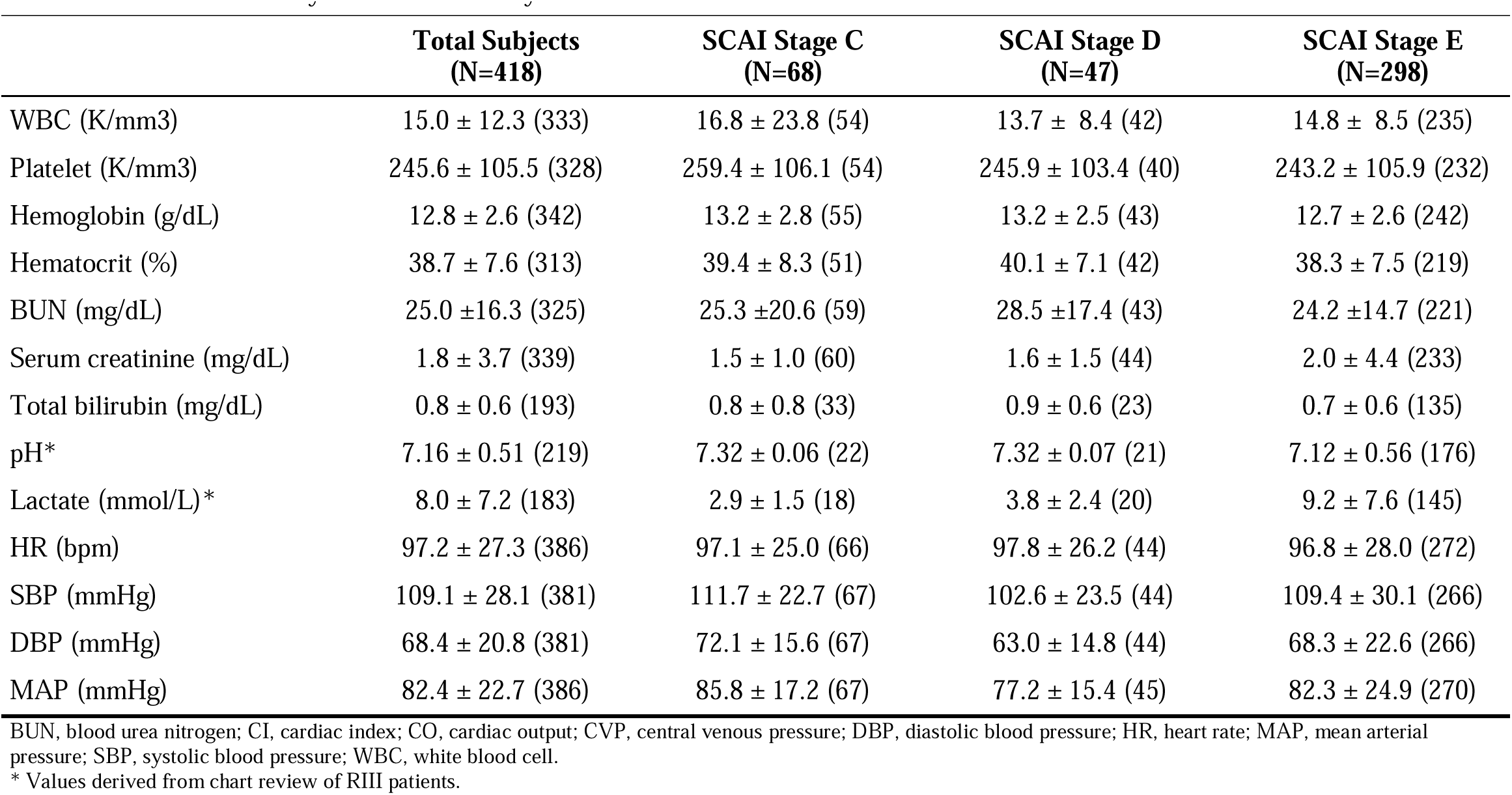
Baseline laboratory values and hemodynamics.

|  | **Total Subjects**<br>**(N=418)** | **SCAI Stage C**<br>**(N=68)** | **SCAI Stage D**<br>**(N=47)** | **SCAI Stage E**<br>**(N=298)** |
| --- | --- | --- | --- | --- |
| WBC (K/mm3) | 15.0 ± 12.3 (333) | 16.8 ± 23.8 (54) | 13.7 ± 8.4 (42) | 14.8 ± 8.5 (235) |
| Platelet (K/mm3) | 245.6 ± 105.5 (328) | 259.4 ± 106.1 (54) | 245.9 ± 103.4 (40) | 243.2 ± 105.9 (232) |
| Hemoglobin (g/dL) | 12.8 ± 2.6 (342) | 13.2 ± 2.8 (55) | 13.2 ± 2.5 (43) | 12.7 ± 2.6 (242) |
| Hematocrit (%) | 38.7 ± 7.6 (313) | 39.4 ± 8.3 (51) | 40.1 ± 7.1 (42) | 38.3 ± 7.5 (219) |
| BUN (mg/dL) | 25.0 ± 16.3 (325) | 25.3 ± 20.6 (59) | 28.5 ± 17.4 (43) | 24.2 ± 14.7 (221) |
| Serum creatinine (mg/dL) | 1.8 ± 3.7 (339) | 1.5 ± 1.0 (60) | 1.6 ± 1.5 (44) | 2.0 ± 4.4 (233) |
| Total bilirubin (mg/dL) | 0.8 ± 0.6 (193) | 0.8 ± 0.8 (33) | 0.9 ± 0.6 (23) | 0.7 ± 0.6 (135) |
| pH* | 7.16 ± 0.51 (219) | 7.32 ± 0.06 (22) | 7.32 ± 0.07 (21) | 7.12 ± 0.56 (176) |
| Lactate (mmol/L)* | 8.0 ± 7.2 (183) | 2.9 ± 1.5 (18) | 3.8 ± 2.4 (20) | 9.2 ± 7.6 (145) |
| HR (bpm) | 97.2 ± 27.3 (386) | 97.1 ± 25.0 (66) | 97.8 ± 26.2 (44) | 96.8 ± 28.0 (272) |
| SBP (mmHg) | 109.1 ± 28.1 (381) | 111.7 ± 22.7 (67) | 102.6 ± 23.5 (44) | 109.4 ± 30.1 (266) |
| DBP (mmHg) | 68.4 ± 20.8 (381) | 72.1 ± 15.6 (67) | 63.0 ± 14.8 (44) | 68.3 ± 22.6 (266) |
| MAP (mmHg) | 82.4 ± 22.7 (386) | 85.8 ± 17.2 (67) | 77.2 ± 15.4 (45) | 82.3 ± 24.9 (270) |

BUN, blood urea nitrogen; CI, cardiac index; CO, cardiac output; CVP, central venous pressure; DBP, diastolic blood pressure; HR, heart rate; MAP, mean arterial pressure; SBP, systolic blood pressure; WBC, white blood cell.
\* Values derived from chart review of RIII patients.

### Admission and procedural characteristics

Admission characteristics are presented in Table 3. Hospital transfers comprised 52.5%, 50.0%, and 41.2% of Stage C, D, and E patients, respectively. All patients experienced acute myocardial infarction, with a majority experiencing STEMI (66.7%, 77.4%, and 80.6% of Stage C, D, and E patients, respectively). Hypoxic brain injury prior to initiation of Impella was observed in 1.6%, 0%, and 9.5% of Stage C, D, and E patients, respectively.

**Table 3.**
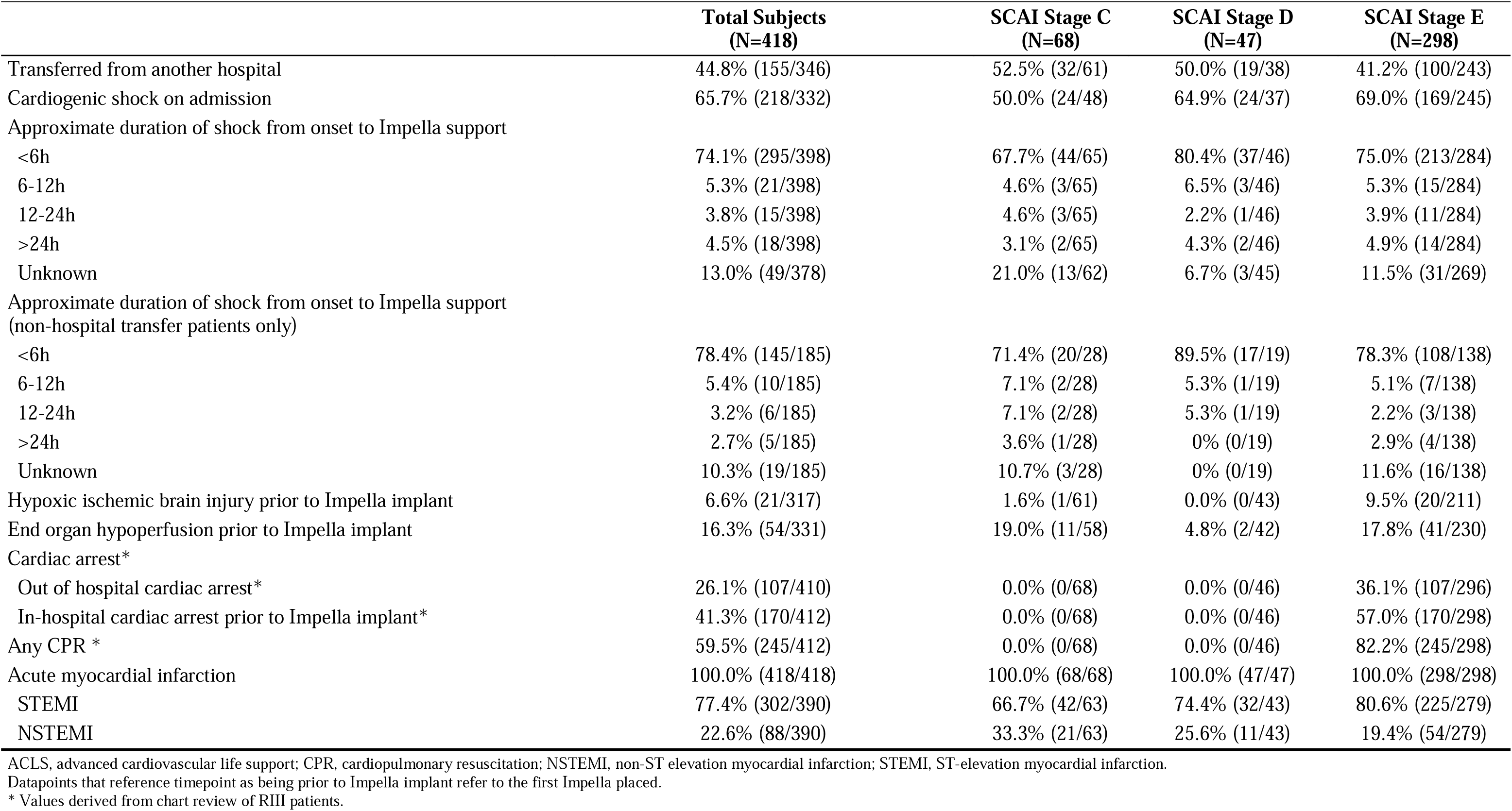
Admission characteristics.

Only 29.4% of Stage C patients were on inotropes/vasopressors prior to initiation of Impella support, whereas 91.5% and 84.8% of Stage D and E patients had received 1 or more inotropes/vasopressors prior to Impella support (Table 4). Relatively few patients received IABP support prior to Impella: 8.8%, 29.8%, and 18.2% of Stage C, D, and E patients, respectively. Extracorporeal membrane oxygenation (ECMO) was performed in 4.3% of Stage E patients prior to Impella. Stage C patients were most likely to have the Impella placed prior to PCI (64.5%) compared to 52.2% in Stage D patients and 56.4% in Stage E patients. A majority of patients underwent pulmonary artery catheterization (86.8%, 82.6%, and 86.1% of Stage C, D, and E patients, respectively).

**Table 4.** Procedural characteristics.

|  | Total Subjects<br>(N=418) | SCAI Stage C<br>(N=68) | SCAI Stage D<br>(N=47) | SCAI Stage E<br>(N=298) |
| --- | --- | --- | --- | --- |
| Patient required inotropes/vasopressors prior to Impella support* | 76.4% (314/411) | 29.4% (20/68) | 91.5% (43/47) | 84.8% (251/296) |
| If yes, maximum number of different inotropes/vasopressors * | 2.1 ± 1.00 (314) | 1.0 ± 0.00 (20) | 1.6 ± 0.49 (43) | 2.2 ± 1.03 (251) |
| MCS (IABP, VAD, ECMO) used prior to Impella implant* | 20.9% (86/412) | 8.8% (6/68) | 29.8% (14/47) | 22.2% (66/297) |
| Patient was supported with an IABP prior to Impella support* | 18.0% (74/412) | 8.8% (6/68) | 29.8% (14/47) | 18.2% (54/297) |
| Duration of index PCI procedure (hours) | 1.7 ± 1.10 (346) | 1.9 ± 1.21 (56) | 1.6 ± 0.94 (42) | 1.7 ± 1.10 (245) |
| Duration of Impella support (hours) | 66.1 ± 65.12 (304) | 64.6 ± 62.34 (46) | 62.8 ± 46.44 (34) | 66.9 ± 68.23 (224) |
| Impella pre PCI | 57.0% (229/402) | 64.5% (40/62) | 52.2% (24/46) | 56.4% (163/289) |
| Door to support time < 90 min | 29.7% (105/354) | 32.8% (19/58) | 17.5% (7/40) | 30.6% (78/255) |
| Door to support time < 90 min (STEMI only) | 38.1% (99/260) | 48.6% (18/37) | 22.2% (6/27) | 37.9% (74/195) |
| Impella CP use | 90.9% (380/418) | 85.3% (58/68) | 95.7% (45/47) | 91.3% (272/298) |
| ICU stay (days) | 10.3 ± 12.65 (351) | 10.3 ± 14.09 (59) | 12.9 ± 18.67 (45) | 9.7 ± 10.72 (245) |
| Duration of index hospitalization (days) | 12.6 ± 13.92 (416) | 12.5 ± 15.24 (67) | 15.1 ± 17.93 (47) | 12.2 ± 12.86 (298) |
| Patients required inotropes/vasopressors during Impella support* | 66.3% (275/415) | 50.7% (34/67) | 57.4% (27/47) | 71.7% (213/297) |
| If yes, maximum number of different inotropes* | 2.2 ± 1.1 (275) | 1.7 ± 0.8 (34) | 2.0 ± 0.9 (27) | 2.3 ± 1.2 (213) |
| Additional devices were implanted post Impella explant* | 14.0% (58/415) | 11.9% (8/67) | 0% (0/47) | 16.2% (48/297) |
| Second Impella implanted | 11.2% (47/418) | 17.6% (12/68) | 12.8% (6/47) | 8.7% (26/298) |
| Impella 2.5/CP | 57.4% (27/47) | 41.7% (5/12) | 66.7% (4/6) | 65.4% (17/26) |
| Impella 5.0/LD | 42.6% (20/47) | 58.3% (7/12) | 33.3% (2/6) | 34.6% (9/26) |
| Contrast volume (ml) | 204.2 ± 105.3 (359) | 198.1 ± 80.21 (59) | 202.7 ± 110.6 (45) | 207.5 ± 109.4 (252) |
| Swan-Ganz/PA Catheter insertion performed | 85.8% (356/415) | 86.8% (59/68) | 82.6% (38/46) | 86.1% (255/296) |

ECMO, extracorporeal membrane oxygenation; IABP, intra-aortic balloon pump; ICU, intensive care unit; MCS, mechanical circulatory support; PA, pulmonary artery; VAD, ventricular assist device.
\* Values derived from chart review of RIII patients.

Lesion characteristics are available in Supplemental Table 1. Stage C/D patients were more likely to have multiple vessels treated than Stage E patients, with 55.0% and 51.1% of Stage C and D patients having 2+ vessels treated, compared to 39.1% of Stage E patients. TIMI flows pre and post PCI were similar across groups, with 86.0%, 82.9%, and 87.0% of Stage C, D, and E patients exhibiting TIMI 3 flow post PCI.

### Clinical outcome

At hospital discharge, MACCE had occurred in 35.3%, 34.0%, and 58.1% of Stage C, D, and E patients, respectively (Table 5). Rate of myocardial infarction and revascularization were similar across SCAI shock classes; however, Stage E patients had a higher rate of stroke/TIA (6.4%) compared to Stage C/D patients (2.9% and 2.1%, respectively), and higher mortality (53.4%) compared to Stage C/D patients (32.4% and 34.0%, respectively). Rates of MACCE at 30 days, 90 days, and 1 year are presented in Table 5 and Supplemental Table 2, respectively. Stage C and E patients had similar length of stay duration (12.5 ± 15.2 days and 12.2 ± 12.9 days, respectively), with a slightly higher length of stay in Stage D patients (15.1 ± 17.9 days).

**Table 5.** In-hospital and 30-day MACCE.

| **Adverse Event** | **Total Subjects**<br>**(N=418)** | **SCAI Stage C**<br>**(N=68 Patients)** | **SCAI Stage D**<br>**(N=47 Patients)** | **SCAI Stage E**<br>**(N=298 Patients)** |
| --- | --- | --- | --- | --- |
| **In-hospital MACCE** | 52.2% (218/418) | 35.3% (24/68) | 34.0% (16/47) | 58.1% (173/298) |
| Death | 48.3% (202/418) | 32.4% (22/68) | 34.0% (16/47) | 53.4% (159/298) |
| Myocardial infarction | 1.2% (5/418) | 1.5% (1/68) | 2.1% (1/47) | 1.0% (3/298) |
| CVA/stroke/TIA | 5.3% (22/418) | 2.9% (2/68) | 2.1% (1/47) | 6.4% (19/298) |
| Revascularization (coronary)/emergent CABG | 1.9% (8/418) | 2.9% (2/68) | 2.1% (1/47) | 1.7% (5/298) |
| **30-day MACCE** | 60.4% (215/356) | 45.3% (24/53) | 44.4% (16/36) | 65.3% (171/262) |
| Death | 55.6% (198/356) | 41.5% (22/53) | 44.4% (16/36) | 59.5% (156/262) |
| Myocardial infarction | 1.4% (5/356) | 1.9% (1/53) | 2.8% (1/36) | 1.2% (3/262) |
| CVA/stroke/TIA | 6.2% (22/356) | 3.8% (2/53) | 2.8% (1/36) | 7.3% (19/262) |
| Revascularization (coronary)/emergent CABG | 2.2% (8/356) | 3.8% (2/53) | 2.8% (1/36) | 1.9% (5/262) |

CVA, cerebrovascular accident; MACCE, major adverse cerebrovascular and cardiac event; TIA, transient ischemic attack.
MACCE through 30 days is a cumulative endpoint, inclusive of in-hospital MACCE events (if occurring before 30 days).

### Change in SCAI shock stage

At baseline SCAI shock stage assessment, 68 patients (16.5%) were classified as Stage C, 47 (11.4%) as Stage D, and 298 (72.2%) as Stage E. A second SCAI classification was performed within 24 hours of Impella support. At ≤ 24 hours, 109 patients (26.4%) were classified as Stage C, 137 patients (33.2%) as Stage D, and 165 patients (40.0%) as Stage E. This represents an absolute 32% improvement in shock severity among Stage E patients within 24 hours of Impella initiation.

Change in SCAI shock stage from baseline assessment to the second assessment performed within 24 hours after Impella initiation is shown in Figure 1. Patients classified as Stage C within 24 hours after Impella initiation showed similarly high in-hospital survival rates regardless of their baseline SCAI SHOCK stage classification (survival rates of 75.9%, 75.0%, and 70.0% in patients with baseline classifications of Stage C, D, and E, respectively). Patients classified as Stage D within 24 hours also showed similar survival, if they were Stage C or D at baseline assessment (survival rates of 69.2% and 68.4%, respectively), though Stage E patients who improved only to Stage D within 24 hours had lower survival (56.5%). Patients classified as Stage E within 24 hours of Impella initiation had poor survival, though survival was lowest in those classified as Stage E at baseline (survival rates of 41.7%, 37.5%, and 31.0%). Patients with Stage E shock at baseline comprised the majority (72%) of the overall RECOVER III population. Of these Stage E patients, 51% showed improvement in SCAI stage within 24 hours after Impella initiation, and 49% remained at Stage E.

**Figure 1.**
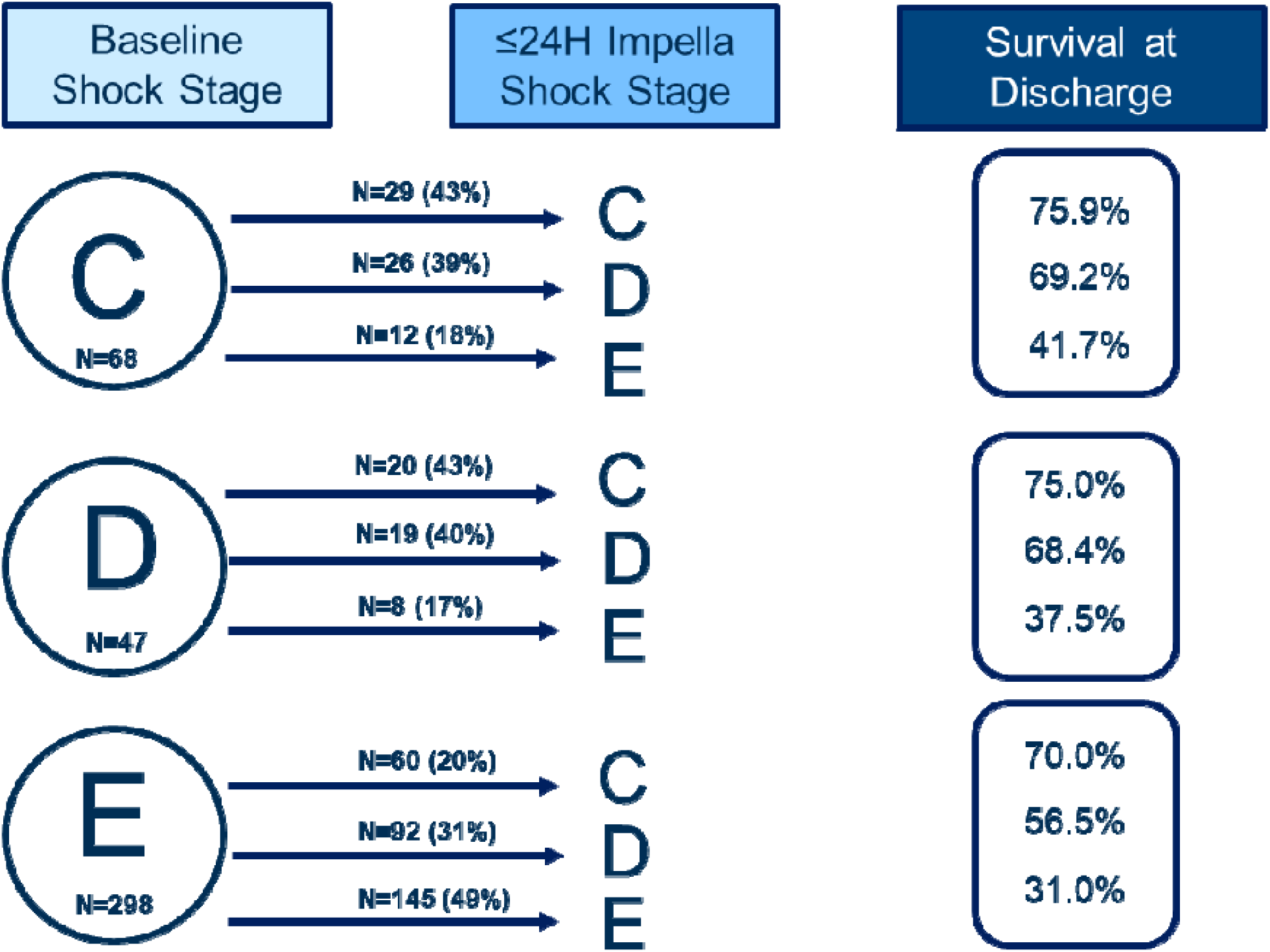
Change in SCAI shock stage from pre Impella to 24 Hours post Impella Initiation. Of 413 patients with SCAI shock stage classification performed prior to Impella implant, 411 had available data to perform repeat SCAI shock stage classification within 24 hours of Impella initiation.

### Kaplan-Meier analyses

Kaplan Meier curve analyses were conducted to assess survival through 30 days according to SCAI assessment of shock stage prior to Impella placement, and within 24 hours after Impella initiation. At 30 days post Impella placement, Kaplan Meier survival estimates in patients classified as Stage C, D, and E at baseline assessment were 59.7%, 56.5%, and 42.9%, respectively (p=0.003; Figure 2). Thirty-day survival estimate in patients classified as Stage C, D, and E within 24 hours after Impella placement were 65.7%, 52.1%, and 29.5%, respectively (p<0.001; Figure 2).

**Figure 2.**
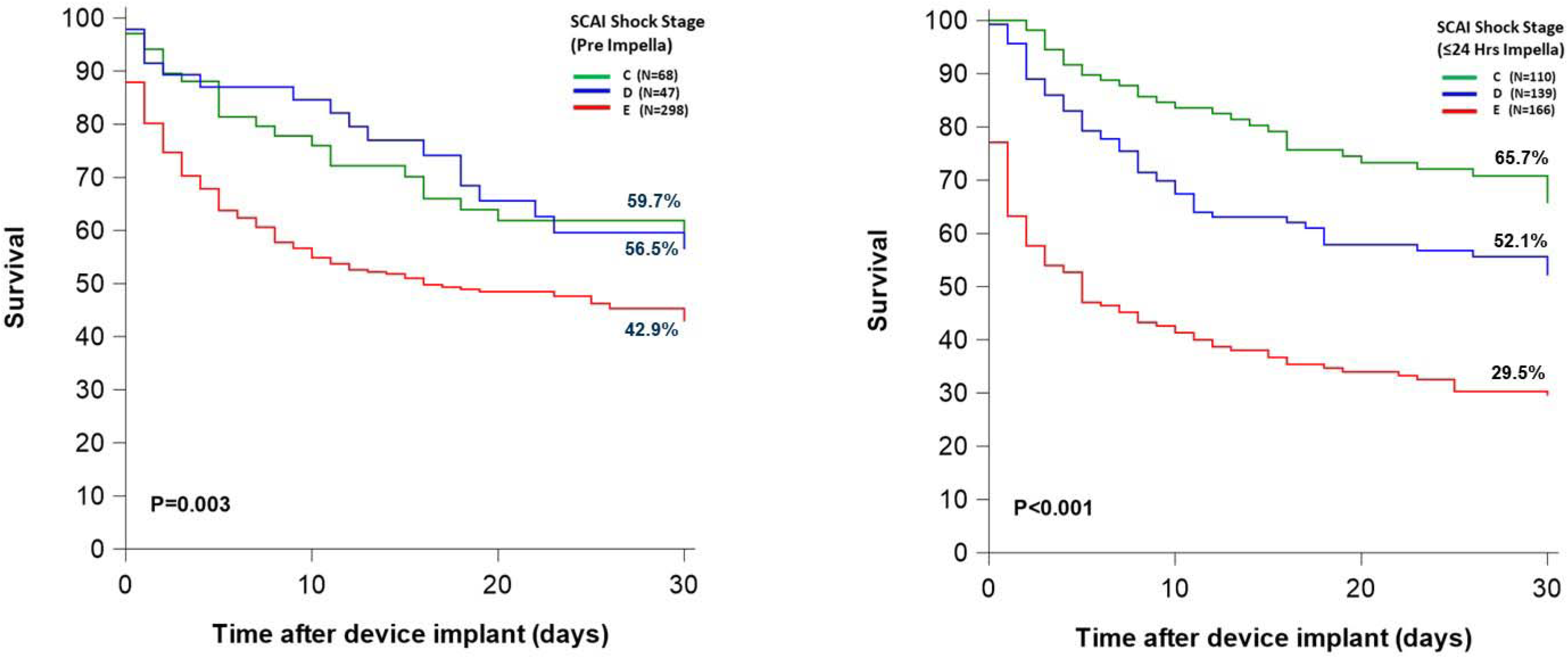
Survival through 30 days. Kaplan Meier survival curve estimates in A) Stage C, D, and E SCAI shock stage subgroups per baseline shock classification (N=68, 47, and 298, respectively); and B) Stage C, D, and E subgroups per classification performed within 24 hours of Impella initiation (N=110, 139, and 166, respectively).

### Predictors of mortality

Regression analyses was performed to assess which of the primary variables used to classify baseline and ≤ 24-hour SCAI shock class were predictive of early mortality. Univariate and multivariate regression analyses is presented in Table 6. In univariate analysis, baseline pH was protective for mortality with each 0.1 increment increase (0.751 odds ratio [OR]; 95% CI, 0.629-0.897; p=0.0016). Increasing lactate was predictive for mortality, though did not attain significance (1.004 OR; 95% CI, 0.998-1.009; p=0.1772). Both pH and lactate were not considered for inclusion in the multivariate model due to a significant number of patients missing data for these variables (only 52% and 44% with available data, respectively).

**Table 6.** Univariate and multivariate regression analysis for 30-day mortality, clinical variables.

| Variable | Univariate |  |  | Multivariate |  |  |
| --- | --- | --- | --- | --- | --- | --- |
|  | Odds Ratio | 95% CI | P value | Odds Ratio | 95% CI | P value |
| IHCA pre Impella | 1.373 | 0.926-2.035 | 0.1146 |  |  |  |
| OHCA pre Impella | 1.292 | 0.831-2.010 | 0.2551 |  |  |  |
| Baseline lactate | 1.004 | 0.998-1.009 | 0.1772 |  |  |  |
| Baseline pH (per 0.1 unit increase) | 0.751 | 0.629-0.897 | 0.0016 |  |  |  |
| No. drugs pre Impella | 1.528 | 1.292-1.807 | <.0001 | 1.356 | 1.124-1.635 | 0.0015 |
| IABP pre Impella | 1.513 | 0.911-2.513 | 0.1094 | 1.699 | 0.964-2.994 | 0.0665 |
| ECMO pre Impella | 0.915 | 0.302-2.771 | 0.8752 |  |  |  |
| Any CPR | 1.673 | 1.124-2.491 | 0.0112 |  |  |  |
| Cardiac arrest post Impella | 12.480 | 5.233-29.764 | <.0001 | 11.078 | 4.534-27.066 | <0.0001 |
| No. drugs post Impella | 1.491 | 1.282-1.734 | <.0001 | 1.343 | 1.136-1.588 | 0.0006 |
| IABP post Impella | 1.055 | 0.211-5.290 | 0.9478 |  |  |  |
| ECMO post Impella | 1.618 | 0.883-2.964 | 0.1195 |  |  |  |

CI, confidence interval. All variables used for classification of baseline and/or 24-hour SCAI SHOCK Stage were included in univariate regression analyses to identify predictors of mortality at 30 days or discharge, whichever is longer. All variables were considered for inclusion in the multivariate model, with the exception of lactate and pH (both with significant missing data, with only 44% and 52% of patients with available baseline lactate and pH values, respectively). A forward fit stepwise approach was taken for multivariate logistic regression, which allowed for forward selection of variables for entry at $p=0.15$ .

In multivariate logistic regression, significant predictors of 30-day mortality were cardiac arrest post Impella placement (11.1 OR; 95% CI, 4.5-27.1; p<0.0001), number of inotropes/vasopressors administered post Impella placement (1.3 OR for each additional drug; 95% CI, 1.1-1.6; p=0.0006), and number of inotropes/vasopressors administered prior to Impella (1.4 OR; 95% CI, 1.1-1.6; p=0.0015). IABP use pre Impella was predictive of mortality, but did not attain statistical significance (1.7 OR; 95% CI, 0.96-3.0; p=0.0665).

A separate logistic regression model assessed the impact of the 6 individual baseline (C, D, or E) and 24-hour SCAI shock stage classifications (C, D, or E) on 30-day mortality (Table 7). A classification of Stage E within 24 hours after Impella placement was the only significant predictor of early mortality (4.8 OR; 95% CI, 2.8-8.3; p<0.001), whereas Stage E classification at baseline was not a significant predictor of mortality (1.5 OR; 95% CI, 0.8-2.6; p=0.186).

**Table 7.** Multivariate analysis for 30-day mortality, SCAI SHOCK stages.

| **Variable** | **Odds Ratio (95% CI)** | **P value** |
| --- | --- | --- |
| Pre Stage C (reference variable) | . | . |
| Pre Stage D | 1.099 (0.489-2.471) | 0.819 |
| Pre Stage E | 1.456 (0.808-2.625) | 0.211 |
| Post Stage C (reference variable) | . | . |
| Post Stage D | 1.625 (0.945-2.795) | 0.079 |
| **Post Stage E** | **4.800 (2.777-8.298)** | **<0.001** |

Pre = SCAI SHOCK Stage classification performed prior to Impella placement. Post = SCAI SHOCK Stage classification performed within 24 hours of Impella placement. A multivariate logistic regression model was run with all SCAI SHOCK stages (from baseline and 24-hour SCAI SHOCK classification) included. The analysis treated each stage as an ordinal staged category, with Pre SCAI Stage C and Post SCAI Stage C the reference variables (i.e., the Post Stage E odds ratio of 4.8 is in relation to the base value/reference variable of Post Stage C.)

## Discussion

In this multi-center, prospective, observational study of AMICS patients who underwent PCI with periprocedural Impella support, results stratified by SCAI shock classification demonstrated several important findings: 1) While a higher baseline shock stage was associated with a higher 30-day mortality rate, SCAI shock stages at ≤24 hours after Impella initiation showed more prognostic value; 2) In patients arriving at Stage E shock and enrolled in RECOVER III, in which all patients received PCI and periprocedural Impella support, improvement in cardiogenic shock stage was noted in 51% of patients; 3) Among the clinical variables used to classify shock stage, only number of vasoactive drugs or cardiac arrest within 24 hours of Impella placement were significantly associated with early mortality.

This is the first large shock study to enroll a predominantly Stage E cohort of patients, with nearly 60% experiencing cardiac arrest prior to Impella placement and an average baseline lactate of 8.0 mmol/L. This is likely due to the fact that this registry exclusively enrolled patients deemed sick enough to require Impella MCS support, whereas other shock registries include patients requiring only lower levels of support (IABP or vasopressors/inotropes only). The comparative utility of Impella versus IABP as therapeutic strategies in this setting has been a source of contention for several years. A recent Medicare claims database analysis comparing LVAD versus other treatments (medical therapy or IABP) for AMICS found that LVAD patients had the highest 30-day mortality; however, the LVAD population was found to have more factors suggesting severe illness.^11^ Furthermore, Almarzooq et al noted that potentially confounding differences in patient and provider factors between treatment groups render any causal inference as to the association between treatment and outcome invalid, pointing to the necessity for randomized controlled trials in this setting in order to perform a legitimate and conclusive comparison of these therapies.^11^

Increased survival of patients presenting at baseline with SCAI Stage E, but improving on Impella to Stage C or D by 24 hours, reinforces the concept that response to therapy may be a more important determinant of survival than the clinical severity of shock at presentation. This observation aligns with prior studies evaluating MCS use in patients with AMICS and empowering physicians to consider this as a “golden 24 hours” of post-shock care.^10, 12, 13^ Similarly, recently published findings from the 11-center Italian Altshock-2 registry found that in cardiogenic shock patients of all etiologies, re-assessment of SCAI shock stage 24 hours after admission provides increased prognostic utility for in-hospital mortality compared to SCAI shock staging performed at time of admission.^14^

The original SCAI shock staging classification was introduced in 2019, with an update in 2022.^10^ Notably, while no prior study evaluating SCAI stages in a shock cohort used exactly the same criteria, many used the original lactate criteria of 5 mmol/L rather than the updated criteria of 8 mmol/L. Recently, the CSWG published detailed evidenced-based criteria helpful in determining shock stage based on their own registry data and clinical outcomes.^10^ The CSWG-SCAI staging criteria had an even higher Stage E criteria of lactate >10 and stipulated that OHCA met criteria for Stage E. We adhered to the CSWG-SCAI criteria in our study; however, Stage E criteria other than lactate and OHCA, including number of vasoactive medications and mechanical circulatory support, frequently played a role. In comparison to prior large shock registries and retrospective studies, the RECOVER III cohort of Stage E patients had similar or worse baseline variables including lactate, pH, vasoactive drug support, and mechanical circulatory support.^10, 12, 13, 15–17^

Interestingly, the 30-day mortality observed in our Stage E patients was comparable or lower than rates seen across published studies, with the exception of NCSI, in which a majority of patients were treated using protocolized best practices.^15^ In the observational, real-world RECOVER III study – a study largely conducted before the publication of NCSI best practices and outcomes – only 11% of patients met all four best practices of right heart catheterization usage, door to support < 90 minutes, pre-PCI Impella, and achieve TIMI 3 flow (Figure 3). In-hospital mortality was 14 percentage points lower in these patients meeting all 4 best practices compared with all other RECOVER III patients (35.6% versus 49.9%).

**Figure 3.**
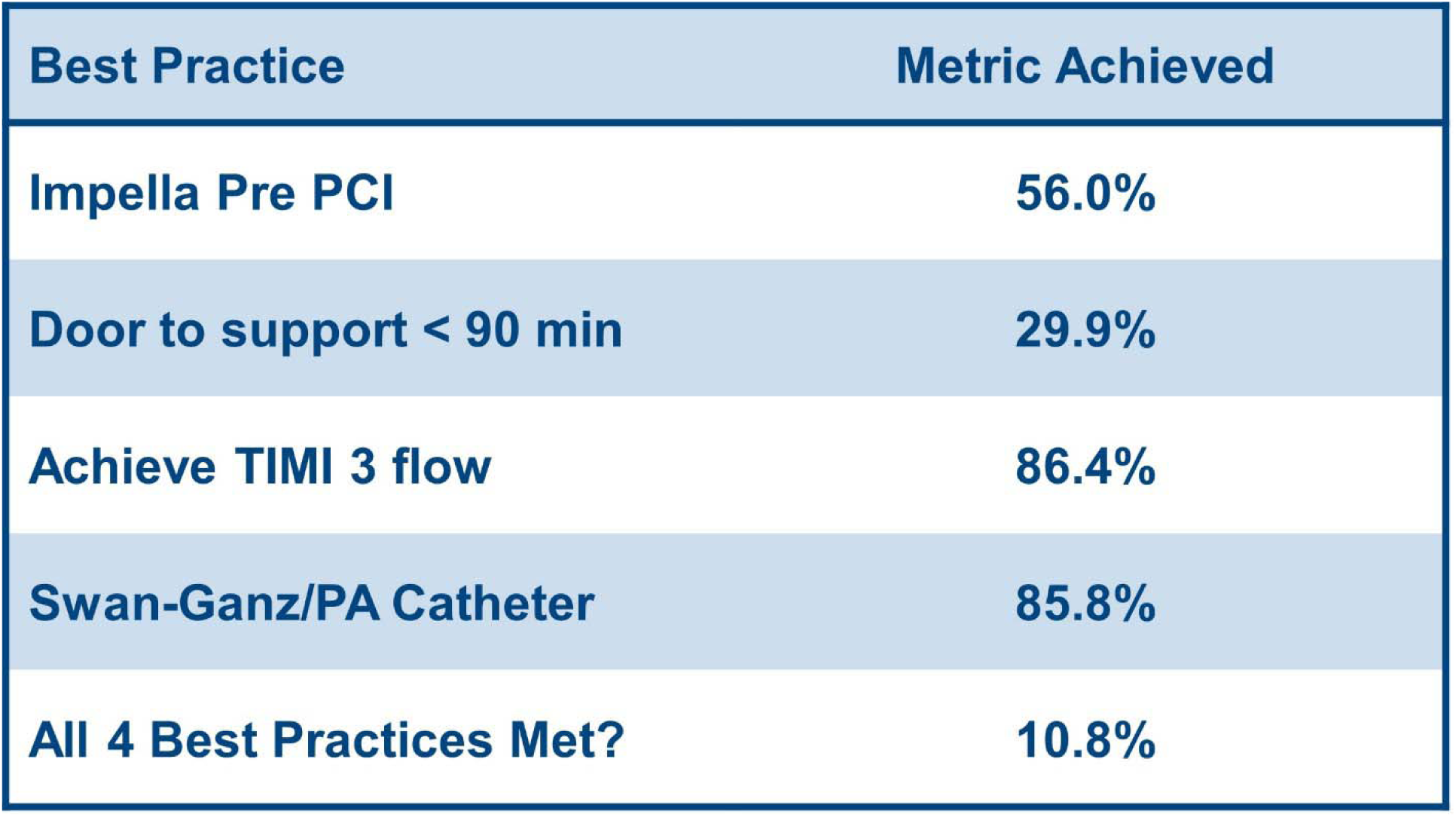
NCSI report card. Utilization of NCSI best practices in the RECOVER III population. Maintain cardiac power output > 0.6W was not included, as a limited number of patients had available data to assess this.

While AMICS “best practices” are actively debated, several publications have described the importance of right heart catheterization usage to guide management. In this “real-world” observational study, 85.8% of patients had a right heart catheterization performed, suggesting that the field may be moving toward universal hemodynamic evaluation in AMICS. Compared to prior cVAD registry published data,^18^ right heart catheterization utilization in AMICS has dramatically increased in the last decade. Nevertheless, only 56% of patients had Impella placement prior to PCI and only 38.1% of STEMI patients (29.9% of all patients) had a “door to support” time less than 90 minutes: far below the metrics achieved by the NCSI study, which protocolized a set of AMICS best practices and achieved a 72% survival to discharge.^7^ The RECOVER IV trial (NCT05506449) will evaluate the use of Impella pre-PCI in STEMI-CS, and the PACCS study (NCT05485376) will evaluate the use of RHC in AMICS. These ongoing randomized controlled trials will hopefully provide additional support for these best practices. Of note, nearly half of the patients in this study were transferred from an outside hospital which speaks to the continued need to establish systems of care in cardiogenic shock patients and hub-and-spoke hospital relationships to ensure appropriate and timely care.^19, 20^

### Study Limitations

There were several limitations to the study. Although the study had a prospective, observational design, there was subsequent retrospective chart review completed to assist in shock classification. Additionally, the shock classifications were done in many cases using an incomplete dataset from varying time points pre-Impella which could introduce staging error given shock staging is dynamic and can rapidly change. Similarly, timing of the second SCAI classification was variable, within the first 24 hours after initiation of Impella support. Notably, a minority of patients had a right heart catheterization done prior to the Impella implantation, making it difficult to ascertain specific hemodynamic metrics at baseline. Finally, the study was unable to assess the utility of best practices in AMICS evaluated (MCS pre-PCI, door to support <90 minutes, TIMI 3 flow, RHC usage, and the maintenance of CPO >0.6, shock team utilization) given the study’s observational design without protocolized or guided management.

## Conclusions

This single-arm, multi-center, prospective, observational study evaluating the use of Impella support in patients with AMICS with predominantly SCAI Stage E at presentation demonstrated an overall 30-day mortality rate of 55.6%. A key finding is that 51% of SCAI Stage E patients at baseline showed SCAI shock stage improvement within 24 hours of Impella support. While baseline shock severity was associated with mortality, SCAI shock stage within 24 hours after Impella initiation showed a more significant association, with Stage E shock at ≤ 24 hours being the only shock stage significantly predictive of 30-day mortality.

## Data Availability

All data produced in the present study are available upon reasonable request to the authors.

## Acknowledgements

Dana Bentley, MWC^®^, provided auxiliary medical writing services and is employed by the device manufacturer.

