## Supplementary material for "Treatment of Acute Myocardial Infarction and Cardiogenic Shock: Outcomes of the RECOVER III Post-Approval Study by SCAI Shock Stage": Figure 1, Tables 1-2

**Supplemental Figure 1. RECOVER III post-approval study flow chart**

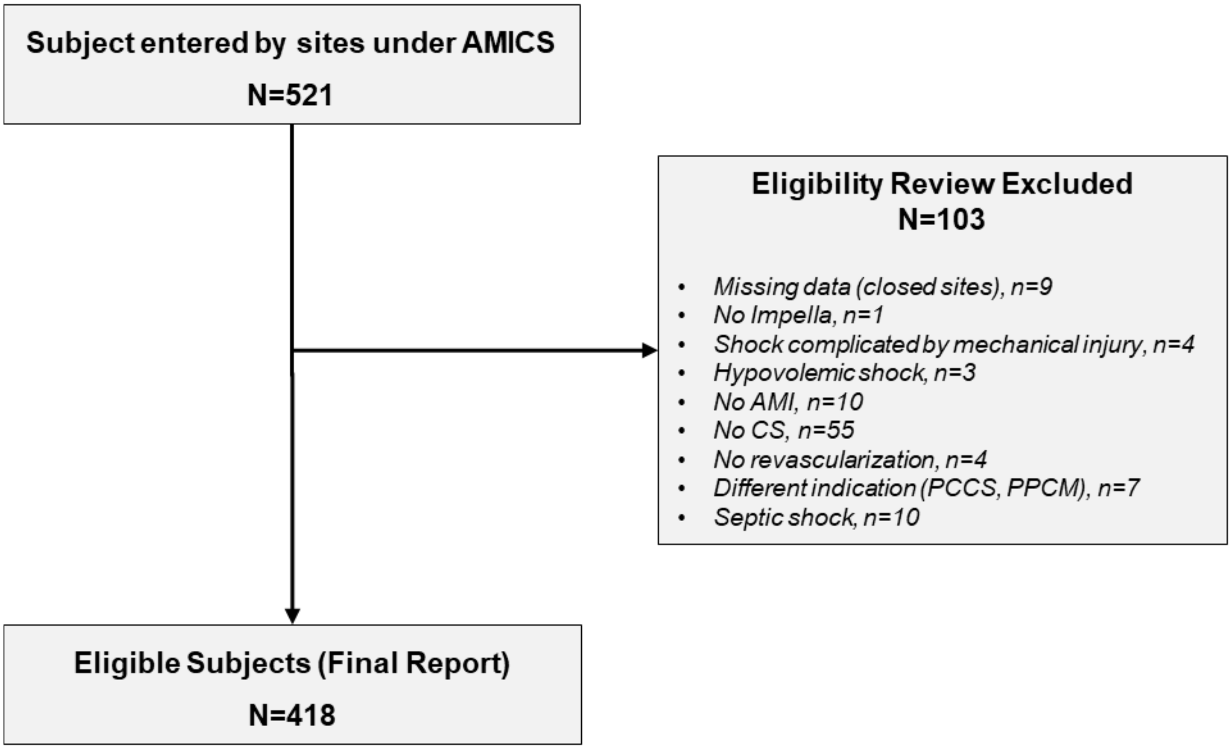

**Supplemental Table 1.** Lesion characteristics, all RECOVER III patients and stratified by baseline SCAI Shock Stage

| | **Total Subjects (N=418)** | **SCAI Stage C (N=68)** | **SCAI Stage D (N=47)** | **SCAI Stage E (N=298)** |
|---|---|---|---|---|
| Number of vessels treated | | | | |
| 1 | 57.2% (219/383) | 45.0% (27/60) | 48.9% (22/45) | 60.9% (167/274) |
| 2 | 29.2% (112/383) | 30.0% (18/60) | 33.3% (15/45) | 28.5% (78/274) |
| 3 | 13.6% (52/383) | 25.0% (15/60) | 17.8% (8/45) | 10.6% (29/274) |
| Target vessel (Patient-based) | | | | |
| GRAFT | 3.1% (12/384) | 3.3% (2/61) | 0.0% ( 0/44) | 3.6% (10/275) |
| LAD | 72.7% (279/384) | 85.2% (52/61) | 70.5% (31/44) | 70.2% (193/275) |
| LCX | 43.0% (165/384) | 59.0% (36/61) | 61.4% (27/44) | 36.7% (101/275) |
| LM | 19.0% (73/384) | 11.5% (7/61) | 25.0% (11/44) | 19.6% (54/275) |
| RCA | 37.0% (142/384) | 45.9% (28/61) | 36.4% (16/44) | 35.6% (98/275) |
| Number of lesions treated | 2.0± 1.27 (382) | 2.3± 1.45 (60) | 2.2± 1.25 (45) | 1.9± 1.22 (273) |
| Target lesion (Patient-based, can have multiple locations) | | | | |
| Distal | 28.0% (106/378) | 21.3% (13/61) | 41.9% (18/43) | 27.4% (74/270) |
| Mid | 55.3% (209/378) | 60.7% (37/61) | 55.8% (24/43) | 53.7% (145/270) |
| Ostial | 8.7% (33/378) | 8.2% (5/61) | 9.3% (4/43) | 8.9% (24/270) |
| Prox | 72.8% (275/378) | 77.0% (47/61) | 74.4% (32/43) | 71.9% (194/270) |
| TIMI flow Pre PCI (minimum) | | | | |
| 0 | 56.1% (162/289) | 57.8% (26/45) | 61.5% (24/39) | 54.7% (111/203) |
| 1 | 7.6% (22/289) | 4.4% (2/45) | 2.6% (1/39) | 9.4% (19/203) |
| 2 | 16.6% (48/289) | 15.6% (7/45) | 12.8% (5/39) | 17.7% (36/203) |
| 3 | 19.7% (57/289) | 22.2% (10/45) | 23.1% (9/39) | 18.2% (37/203) |
| TIMI flow Post PCI (minimum) | | | | |
| 0 | 4.1% (13/317) | 4.0% (2/50) | 7.3% (3/41) | 3.6% (8/224) |
| 1 | 1.3% (4/317) | 2.0% (1/50) | 0% (0/41) | 1.3% (3/224) |
| 2 | 8.2% (26/317) | 8.0% (4/50) | 9.8% (4/41) | 8.0% (18/224) |
| 3 | 86.4% (274/317) | 86.0% (43/50) | 82.9% (34/41) | 87.1% (195/224) |

LAD, left anterior descending; LCX, left circumflex; LM, left main; RCA, right coronary artery.
Values are expressed as percentage (numerator/denominator).

**Supplemental Table 2.** 90-day and 1-year MACCE

| Adverse Event | Total Subjects (N=418) | SCAI Stage C (N=68 Patients) | SCAI Stage D (N=47 Patients) | SCAI Stage E (N=298 Patients) |
|---|---|---|---|---|
| **90-day MACCE** | 65.7% (222/338) | 50.0% (26/52) | 51.5% (17/33) | 70.2% (174/248) |
| Death | 60.7% (205/338) | 44.2% (23/52) | 51.5% (17/33) | 64.5% (160/248) |
| Myocardial infarction | 2.1% (7/338) | 3.8% (2/52) | 3.0% (1/33) | 1.6% (4/248) |
| CVA/stroke/TIA | 6.5% (22/338) | 3.8% (2/52) | 3.0% (1/33) | 7.7% (19/248) |
| Revascularization (coronary)/emergent CABG | 2.7% (9/338) | 3.8% (2/52) | 3.0% (1/33) | 2.4% (6/248) |
| **1-year MACCE** | 71.4% (230/322) | 54.9% (28/51) | 62.1% (18/29) | 75.5% (179/237) |
| Death | 65.2% (210/322) | 45.1% (23/51) | 58.6% (17/29) | 69.6% (165/237) |
| Myocardial infarction | 3.1% (10/322) | 3.9% (2/51) | 6.9% (2/29) | 2.5% (6/237) |
| CVA/stroke/TIA | 7.1% (23/322) | 5.9% (3/51) | 3.4% (1/29) | 8.0% (19/237) |
| Revascularization (coronary)/emergent CABG | 3.4% (11/322) | 7.8% (4/51) | 3.4% (1/29) | 2.5% (6/237) |

MACCE through 90 days and 1 year are cumulative endpoints.
